# Racial and Ethnic Disparities in the Utilization and Outcomes with WATCHMAN FLX: A SURPASS analysis of the NCDR LAAO Registry

**DOI:** 10.1101/2024.05.06.24306969

**Authors:** Oluseun O Alli, Jalaj Garg, Brian C. Boursiquot, Samir R Kapadia, Robert W Yeh, Matthew J Price, Jonathan P Piccini, Devi G Nair, Jonathan C Hsu, Douglas N Gibson, TDominic Alloco, Thomas Christen, Brad Sutton, James V Freeman

## Abstract

**Background:** Left atrial appendage occlusion (LAAO) is increasingly used as an alternative to oral anticoagulation for stroke prevention in select patients with atrial fibrillation. Data on outcomes in racial and ethnic minority individuals are limited. This analysis assessed differences in the utilization and outcomes of LAAO by race and ethnicity in a large national registry.

**Methods:** This analysis acquired data on WATCHMAN FLX patients from the National Cardiovascular Data Registry (NCDR) LAAO Registry through September 2022. All patients with an attempted WATCHMAN FLX implantation and known race and ethnicity were included. Baseline characteristics and 1 year event rates were compared.

**Results:** A total of 97,185 patients were analyzed; 87,339 were White (90%), 3,750 Black (Black/African American 3.9%), and 2,866 Hispanic ([Hispanic/Latinx] 2.9%). Black and Hispanic patients were younger with a higher incidence of prior stroke and significant bleeding compared to White patients. Black and Hispanic patients were treated with LAAO in smaller numbers relative to their proportion of the US population. Rates of procedural success were similar between groups. Though direct oral anticoagulants were prescribed in most patients across the groups, dual and single antiplatelet therapy were prescribed more often in Black patients. Black patients had significantly higher rates of 1-year death and bleeding compared to White and Hispanic patients.

**Conclusions:** Patients from racial and ethnic minority groups comprise a disproportionately small fraction of all patients who undergo LAAO. Black and Hispanic patients were younger but had significantly higher comorbidities compared to White patients. Procedural success was similar amongst the groups but Black patients experienced higher rates of death and bleeding at 1 year.

**Graphic Abstract:** 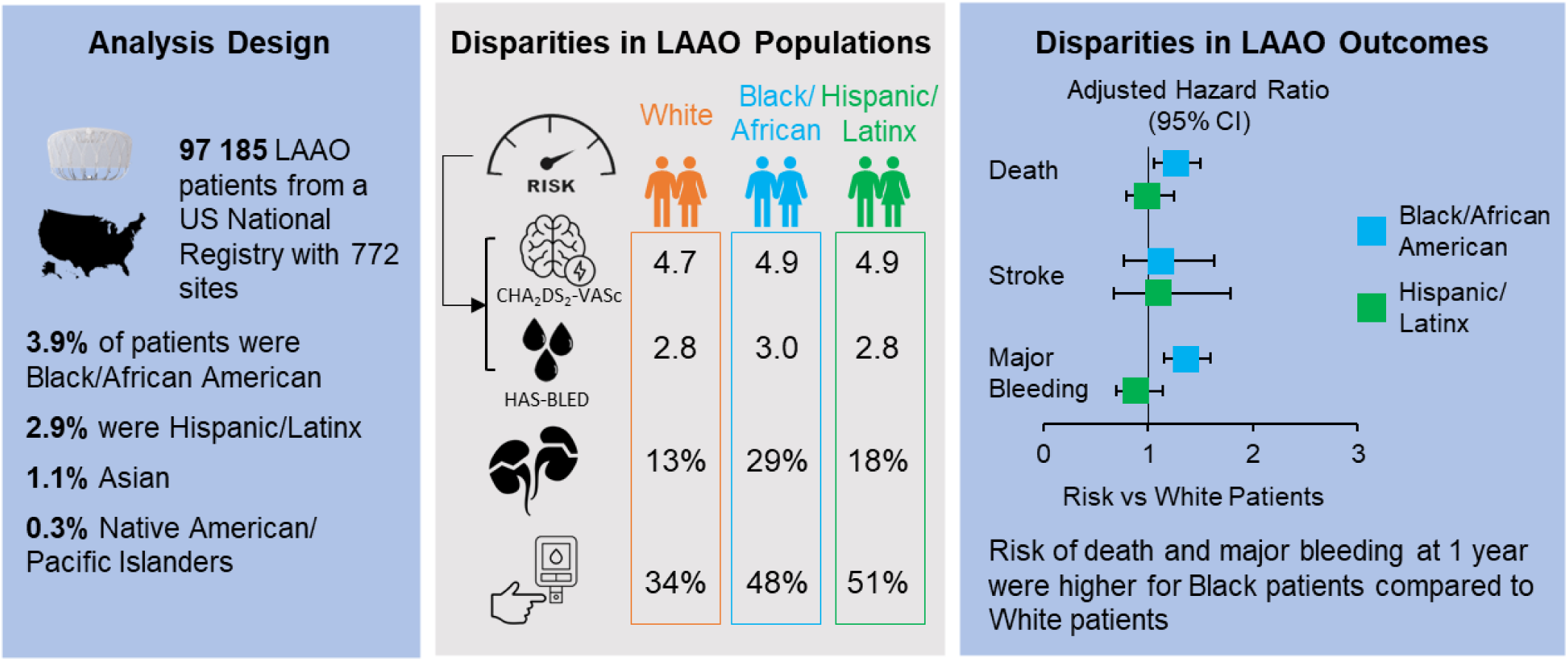

**What is known:** There is underutilization of LAAO among racial and ethnic minority patients with AF and there are racial and ethnic differences in periprocedural outcomes.

**What this study adds:** In this study from a large US national registry of patients undergoing LAAO, Black patients were younger but had higher baseline comorbidities and experienced higher rates of bleeding at 45 days and 1-year and higher 1-year mortality. Further work is needed to enroll diverse patients into research trials and to provide equitable AF-related access to advanced care and intra- procedural and post-procedural care in US real-world practice.

## Introduction

Transcatheter left atrial appendage occlusion (LAAO) is an increasingly utilized alternative therapy to long-term anticoagulation for stroke prevention in selected patients with nonvalvular atrial fibrillation (AF). Stroke is the most devastating complication of AF, and the use of oral anticoagulation or LAAO has been shown to be effective at reducing stroke risk.^1,2^ Despite the effectiveness of anticoagulation, racial and ethnic minority patients, and particularly Black patients diagnosed with AF, are less likely to be discharged on oral anticoagulants, while also having a higher risk of stroke compared to White patients.^3,4^ Similarly, it has been shown that there are significant racial and ethnic disparities in structural heart disease interventions, including LAAO^5^, with one study showing that racial and ethnic minority patients comprised less than 4% of those treated with these advanced interventions between 2011 and 2016.^5^ Racial and ethnic minority individuals have also been consistently shown to be undertreated in terms of other cardiovascular procedures like transcatheter aortic valve replacement (TAVR), mitral valve replacement, and implantation of implantable cardioverter-defibrillators.^6–9^

In published clinical trials and real-world experience with transcatheter LAAO, non- Hispanic White patients have represented over 90% of cases; it is not clear that the efficacy in the overall population is generalizable to all racial and ethnic groups. Studies of racial and ethnic disparities in transcatheter LAAO outcomes have shown higher rates of in-hospital complications among non-Hispanic Black and Hispanic patients compared to non-Hispanic White patients.^10^ However, there is a paucity of data comparing post-discharge outcomes across racial and ethnic groups.

The SURPASS registry is an analysis of patients who underwent LAAO with the WATCHMAN FLX device as part of the American College of Cardiology (ACC) Foundation’s National Cardiovascular Data Registry (NCDR) LAAO Registry. The aim of this current analysis is to examine the utilization of and outcomes after WATCHMAN FLX LAAO across racial and ethnic groups in a large national registry.

## Methods

### Data Source

This analysis included data on patients implanted with the WATCHMAN FLX device (Boston Scientific, Marlborough, MA, USA) and included in the NCDR LAAO Registry. This registry was initiated in December 2015 after FDA approval of the first WATCHMAN device.^11^ The NCDR LAAO Registry serves as the formal post-market surveillance vehicle required by the FDA for the WATCHMAN device, and it is the only registry approved by CMS to satisfy the coverage decision data submission requirement.^12^ As of April 2016, U.S. hospitals were required to submit data for all WATCHMAN procedures into the LAAO Registry to qualify for Medicare reimbursement. Hospitals are encouraged to submit data on all device recipients regardless of insurance status.

The LAAO Registry data collection methods have been detailed previously.^11^ In brief, the LAAO Registry collects approximately 220 data elements from the implant hospitalization, 60 for each follow-up visit, and 15 data elements to support the adjudication of adverse events. Data are collected at discharge, and follow-up visits over the first year occur at 45 days (±14 days), 180 days (-30 days/+60 days), and 365 days (±60 days). The NCDR Data Quality Reporting process is designed to ensure that submissions are complete, valid, and accurate; it involves an annual audit of about 5% of sites that are randomly selected during which submitted data are compared with source documentation and billing data.^13^ The LAAO Registry developed and validated a novel process to adjudicate adverse clinical events over follow-up. A computer-based algorithm uses discrete combinations of registry data elements based on standard event definitions to adjudicate adverse events.^14^ Cases are manually adjudicated when registry data elements are incomplete or conflicting. Adjudicated adverse events in the registry include ischemic stroke, hemorrhagic stroke, undetermined stroke, transient ischemic attack, intracranial hemorrhage, systemic arterial embolism (other than stroke), major bleeding, and major vascular complications.

### Study Population

This analysis utilized patient data collected between August 5, 2020 to September 30, 2022. All patients with attempted WATCHMAN FLX implantation were included. The data collection form of the NCDR LAAO registry has a racial category question and a Hispanic or Latinx ethnicity question. Analysis cohorts were based on the selections within these two questions. Race and/or Hispanic or Latinx ethnicity was determined by the patient/family and site-reported. Groups were then defined as White [non-Hispanic White], Black [non-Hispanic Black/African American], Asian [non-Hispanic Asian], Native American/Pacific Islander [non-Hispanic American Indian/Alaska Native/Native Hawaiian/Pacific Islander] and Hispanic [Hispanic or Latinx]. Additional groups included: multiple selections (if multiple categories were selected), other (one of the two questions was not answered), and no race or ethnicity reported (neither question was answered). Patient and procedure characteristics and outcomes in patients with multiple selections, other or no selection can be found in **Supplemental Tables 1-4**.

### Study Endpoints

All-cause death, stroke [ischemic and/or hemorrhagic], and major bleeding [any bleeding requiring hospitalization, and/or causing a decrease in hemoglobin level > 2g/dL, and/or requiring blood transfusion that was not hemorrhagic stroke ^11^] were evaluated at 45 days and 1 year. Additional endpoints evaluated at 45 days include major vascular complications, pericardial effusion, device embolization, device migration, and peridevice leak.

### Statistical Analysis

Data were summarized using descriptive statistics for continuous variables and frequency tables or proportions for discrete variables. P-values were from the Fisher exact or chi square test, as appropriate. Clinical event rates at 45 days and 1 year were estimated using the Kaplan-Meier method with p-values from the log-rank test. Adjustment or differences in baseline characteristics was performed in the three largest groups. Cox proportional hazards regression was performed for the comparisons between White and Black and White vs Hispanic patients.

Candidate variables are included in the **Supplement**. The following variables, based on clinical relevance and with a p<0.02 in univariate analyses were included: age, gender, components of the CHA_2_DS_2_-VASc and/or HAS-BLED scores, type of AF, fall risk, chronic lung disease, sleep apnea, cardiomyopathy, coronary artery disease, prior ablation, and left ventricular ejection fraction. Statistical analyses were conducted using SAS version 9.4 (SAS Institute, Cary, North Carolina).

## Results

A total of 97,185 patients were implanted with WATCHMAN FLX between August 5, 2020 and September 30, 2022 at 772 sites in the United States. Most patients were White (90%, n=87339); Black patients comprised 3.9% (n=3750), Hispanic 2.9% (n=2866), Asian 1.1% (n=1082), and American Indian/Pacific Islander 0.3% (n=274) of the patient population. Data from other groups can be found in the **Supplement** (**Supplemental Tables 1-3, 5 and 7**) including 297 patients with multiple races/ethnicities selected (0.3%), 1255 (1.3%) who did not have data for both race and ethnicity and 322 who did not have data for either race or ethnicity (0.3%).

### Baseline Characteristics

Overall, there were significant differences in baseline characteristics between racial and ethnic groups. Black and Native American/Pacific Islander patients were several years younger than other groups and had the largest ratio of women to men (**Table 1**). The CHA_2_DS_2_-VASc score was highest in Black, Hispanic and Native American/Pacific Islander patients; and HAS-BLED was highest in Black patients. Prior stroke was highest in Black and Asian patients and prior bleeding highest in Black patients.

**Table 1.** Baseline Characteristics.

| **Description** | **White**<br>**(N = 87339)** | **Black/African**<br>**American**<br>**(N = 3750)** | **Hispanic/**<br>**Latinx**<br>**Ethnicity**<br>**(N = 2866)** | **Asian**<br>**(N = 1082)** | **American Indian/**<br>**Alaska Native or**<br>**Native Hawaiian/**<br>**Pacific Islander**<br>**(N = 274)** |
| --- | --- | --- | --- | --- | --- |
| Age, years | 76.6 ± 7.6<br>(87339) | 72.3 ± 9.5 (3750) | 75.0 ± 9.1<br>(2866) | 75.5 ± 9.0<br>(1082) | 73.5 ± 9.1 (274) |
| Age group |  |  |  |  |  |
| <70 | 14958 (17.1%) | 1352 (36.1%) | 698 (24.4%) | 263 (24.3%) | 85 (31.0%) |
| 70-80 | 44684 (51.2%) | 1658 (44.2%) | 1332 (46.5%) | 483 (44.6%) | 129 (47.1%) |
| >80 | 27697 (31.7%) | 740 (19.7%) | 836 (29.2%) | 336 (31.1%) | 60 (21.9%) |
| Female | 35762 (41.0%) | 1834 (48.9%) | 1240 (43.3%) | 443 (40.9%) | 120 (43.8%) |
| CHA<sub>2</sub>DS<sub>2</sub>-VASc Score | 4.7 ± 1.5 (87088) | 4.9 ± 1.6 (3745) | 4.9 ± 1.6 (2861) | 4.8 ± 1.5 (1077) | 4.9 ± 1.5 (274) |
| Congestive heart failure | 32477 (37.2%) | 1835 (49.0%) | 1101 (38.4%) | 347 (32.1%) | 116 (42.3%) |
| LV dysfunction | 7471 (8.6%) | 528 (14.1%) | 310 (10.8%) | 77 (7.1%) | 28 (10.2%) |
| Hypertension | 79721 (91.3%) | 3597 (95.9%) | 2689 (93.8%) | 1000 (92.6%) | 256 (93.4%) |
| Diabetes Mellitus | 29429 (33.7%) | 1775 (47.4%) | 1462 (51.0%) | 508 (47.0%) | 122 (44.5%) |
| Stroke | 17298 (19.8%) | 1134 (30.3%) | 760 (26.5%) | 343 (31.8%) | 73 (26.6%) |
| Transient ischemic attack | 10015 (11.5%) | 377 (10.1%) | 297 (10.4%) | 108 (10.0%) | 44 (16.1%) |
| Thromboembolic event | 12183 (14.0%) | 738 (19.7%) | 470 (16.4%) | 183 (16.9%) | 44 (16.1%) |
| Vascular disease | 46108 (52.8%) | 1808 (48.2%) | 1394 (48.7%) | 491 (45.5%) | 148 (54.0%) |
| HAS-BLED Score | 2.8 ± 1.1 (87081) | 3.0 ± 1.2 (3742) | 2.8 ± 1.2 (2861) | 2.8 ± 1.1 (1077) | 2.8 ± 1.2 (273) |
| Uncontrolled hypertension | 25081 (28.7%) | 1314 (35.1%) | 904 (31.5%) | 270 (25.0%) | 70 (25.6%) |
| Abnormal renal function | 11315 (13.0%) | 1079 (28.8%) | 495 (17.3%) | 210 (19.4%) | 51 (18.7%) |
| Abnormal liver function | 2598 (3.0%) | 136 (3.6%) | 148 (5.2%) | 39 (3.6%) | 14 (5.1%) |
| Stroke | 18051 (20.7%) | 1162 (31.0%) | 774 (27.0%) | 361 (33.4%) | 82 (30.0%) |
| Bleeding | 56949 (65.3%) | 2829 (75.5%) | 1929 (67.3%) | 745 (69.0%) | 194 (71.1%) |
| Labile INR | 4277 (4.9%) | 173 (4.6%) | 145 (5.1%) | 35 (3.2%) | 14 (5.1%) |
| Alcohol | 4415 (5.1%) | 145 (3.9%) | 92 (3.2%) | 26 (2.4%) | 14 (5.1%) |
| Drugs – Antiplatelet | 29970 (34.4%) | 1325 (35.4%) | 879 (30.7%) | 305 (28.2%) | 101 (37.0%) |
| Drugs – NSAIDS | 11418 (13.1%) | 498 (13.3%) | 288 (10.1%) | 106 (9.8%) | 24 (8.8%) |
| Increased Fall Risk | 36969 (42.4%) | 1256 (33.6%) | 1187 (41.5%) | 394 (36.5%) | 109 (39.9%) |
| Clinically relevant bleeding event | 48158 (55.3%) | 2712 (72.4%) | 1732 (60.5%) | 690 (63.8%) | 162 (59.1%) |
| Atrial fibrillation pattern |  |  |  |  |  |
| Paroxysmal | 53806 (62.1%) | 2598 (69.9%) | 1782 (62.9%) | 635 (59.2%) | 182 (66.9%) |
| Persistent | 18019 (20.8%) | 640 (17.2%) | 568 (20.0%) | 217 (20.2%) | 47 (17.3%) |
| Long-standing persistent | 10189 (11.8%) | 322 (8.7%) | 312 (11.0%) | 137 (12.8%) | 30 (11.0%) |
| Permanent | 4677 (5.4%) | 159 (4.3%) | 173 (6.1%) | 84 (7.8%) | 13 (4.8%) |
| Attempt at AF termination of patients | 40546 (46.5%) | 1325 (35.4%) | 1018 (35.6%) | 368 (34.0%) | 102 (37.2%) |
| with AF |  |  |  |  |  |
| Atrial flutter | 15980 (18.3%) | 720 (19.2%) | 425 (14.9%) | 163 (15.1%) | 46 (16.8%) |
| Attempt at atrial flutter termination of<br>patients with atrial flutter | 10834 (68.2%) | 436 (60.8%) | 282 (66.8%) | 103 (63.2%) | 34 (73.9%) |
| Cardiac structural intervention | 6824 (7.8%) | 240 (6.4%) | 218 (7.6%) | 73 (6.8%) | 18 (6.6%) |
| Hypertrophic cardiomyopathy | 880 (1.0%) | 70 (1.9%) | 25 (0.9%) | 15 (1.4%) | 4 (1.5%) |
| Chronic lung disease | 17303 (19.8%) | 799 (21.3%) | 415 (14.5%) | 115 (10.6%) | 67 (24.5%) |
| Coronary artery disease | 38750 (44.4%) | 1464 (39.1%) | 1183 (41.4%) | 429 (39.7%) | 126 (46.0%) |
| Sleep apnea | 26587 (30.5%) | 1071 (28.6%) | 667 (23.3%) | 195 (18.0%) | 86 (31.4%) |
| Sleep apnea treatment | 19489 (22.5%) | 727 (19.6%) | 434 (15.3%) | 139 (12.9%) | 61 (22.3%) |
| LAA orifice maximal width, mm | 20.9 ± 4.3<br>(85401) | 22.0 ± 4.5 (3683) | 20.7 ± 4.4<br>(2797) | 21.5 ± 4.3<br>(1064) | 21.2 ± 4.7 (270) |
| Left ventricular ejection fraction, % | 54.3 ± 9.6<br>(56095) | 52.5 ± 12.1 (2532) | 53.7 ± 10.8<br>(1801) | 55.7 ± 10.0<br>(668) | 53.5 ± 10.6 (185) |

Numbers are n (%) or Mean±SD (N); Abbreviations: AF=atrial fibrillation; INR=international normalized ratio; NSAIDS=Non-steroidal anti-inflammatory drugs

### Procedural Characteristics

Indications for LAAO were consistent across the groups and mainly related to risk of bleeding and/or stroke (**Table 2**). Device size tended to be larger in Black and Asian patients. Same day discharge rates ranged from 21.5% to 38.2%. The rates of successful implantation and complete seal at post-procedure were high in all groups. Direct oral anticoagulation (DOAC) alone and DOAC + aspirin were prescribed in the majority of patients across all groups. Dual antiplatelet therapy (DAPT) and single antiplatelet therapy (SAPT) were prescribed more often in Black patients than other groups (**Table 3**).

**Table 2.** Procedural Characteristics.

| **Description** | **White**<br>**(N = 87339)** | **Black/African**<br>**American**<br>**(N = 3750)** | **Hispanic/**<br>**Latinx**<br>**Ethnicity**<br>**(N = 2866)** | **Asian**<br>**(N = 1082)** | **American Indian/**<br>**Alaska Native or**<br>**Native Hawaiian/**<br>**Pacific Islander**<br>**(N = 274)** |
| --- | --- | --- | --- | --- | --- |
| Successful implantation | 85149 (97.5%) | 3688 (98.4%) | 2797 (97.6%) | 1053 (97.3%) | 264 (96.4%) |
| Indication for procedure* |  |  |  |  |  |
| Increased thromboembolic stroke risk | 55482 (63.6%) | 2348 (62.6%) | 1844 (64.3%) | 723 (66.8%) | 182 (66.7%) |
| History of major bleed | 44915 (51.4%) | 2562 (68.3%) | 1612 (56.3%) | 631 (58.3%) | 151 (55.3%) |
| High fall risk | 33484 (38.4%) | 1116 (29.8%) | 1081 (37.7%) | 359 (33.2%) | 95 (34.8%) |
| Labile INR | 3200 (3.7%) | 145 (3.9%) | 109 (3.8%) | 34 (3.1%) | 7 (2.6%) |
| Patient preference | 37599 (43.1%) | 1322 (35.3%) | 1118 (39.0%) | 479 (44.3%) | 117 (42.9%) |
| Non-compliance with anticoagulation<br>therapy | 2806 (3.2%) | 145 (3.9%) | 128 (4.5%) | 23 (2.1%) | 12 (4.4%) |
| Average number of devices used per<br>patient | 1.2 ± 0.5 (87339) | 1.1 ± 0.4 (3750) | 1.1 ± 0.4 (2866) | 1.2 ± 0.4 (1082) | 1.2 ± 0.55 (274) |
| Device size, mm |  |  |  |  |  |
| 20 | 10604 (12.5%) | 253 (6.9%) | 340 (12.2%) | 79 (7.5%) | 28 (10.6%) |
| 24 | 26669 (31.3%) | 859 (23.3%) | 915 (32.7%) | 279 (26.5%) | 75 (28.4%) |
| 27 | 27026 (31.7%) | 1242 (33.7%) | 875 (31.3%) | 360 (34.2%) | 85 (32.2%) |
| 31 | 15163 (17.8%) | 898 (24.4%) | 475 (17.0%) | 212 (20.1%) | 54 (20.5%) |
| 35 | 5687 (6.7%) | 436 (11.8%) | 192 (6.9%) | 123 (11.7%) | 22 (8.3%) |
| Procedure time, minutes | 78.0 ± 66.2<br>(87336) | 84.9 ± 57.5 (3750) | 88.9 ± 274.1<br>(2866) | 84.7 ± 86.6<br>(1081) | 75.2 ± 37.2 (274) |
| Contrast volume, mL | 37.9 ± 33.2<br>(84440) | 36.5 ± 33.9 (3558) | 43.6 ± 37.1<br>(2736) | 38.9 ± 34.8<br>(1039) | 40.6 ± 31.2 (267) |
| General anesthesia | 82633 (94.8%) | 3544 (94.8%) | 2736 (95.8%) | 1033 (95.8%) | 265 (96.7%) |
| Concomitant procedure performed | 1776 (2.0%) | 86 (2.3%) | 59 (2.1%) | 18 (1.7%) | 8 (2.9%) |
| Length of stay, days | 2.0 ± 6.76<br>(87339) | 2.6 ± 9.05 (3750) | 2.6 ± 10.19<br>(2866) | 2.2 ± 2.75<br>(1082) | 1.8 ± 1.19 (274) |
| Same day discharge | 30301 (34.7%) | 940 (25.1%) | 652 (22.8%) | 287 (26.5%) | 114 (41.6%) |
| Residual leak post procedure |  |  |  |  |  |
| 0 mm | 81155 (96.4%) | 3466 (94.8%) | 2661 (96.7%) | 980 (94.6%) | 242 (94.5%) |
| >0 mm to ≤3 mm | 2414 (2.9%) | 165 (4.5%) | 77 (2.8%) | 43 (4.2%) | 11 (4.3%) |
| >3 mm to ≤5 mm | 571 (0.7%) | 24 (0.7%) | 13 (0.5%) | 11 (1.1%) | 3 (1.2%) |
| >5 mm | 19 (0%) | 1 (0.03%) | 0 (0%) | 2 (0.2%) | 0 (0%) |
| Patients with missing leak data | 3180 | 94 | 115 | 46 | 18 |

Numbers are n (%) or Mean±SD (N); \*patients can have more than one indication; Abbreviations: INR=international normalized ratio

**Table 3.**
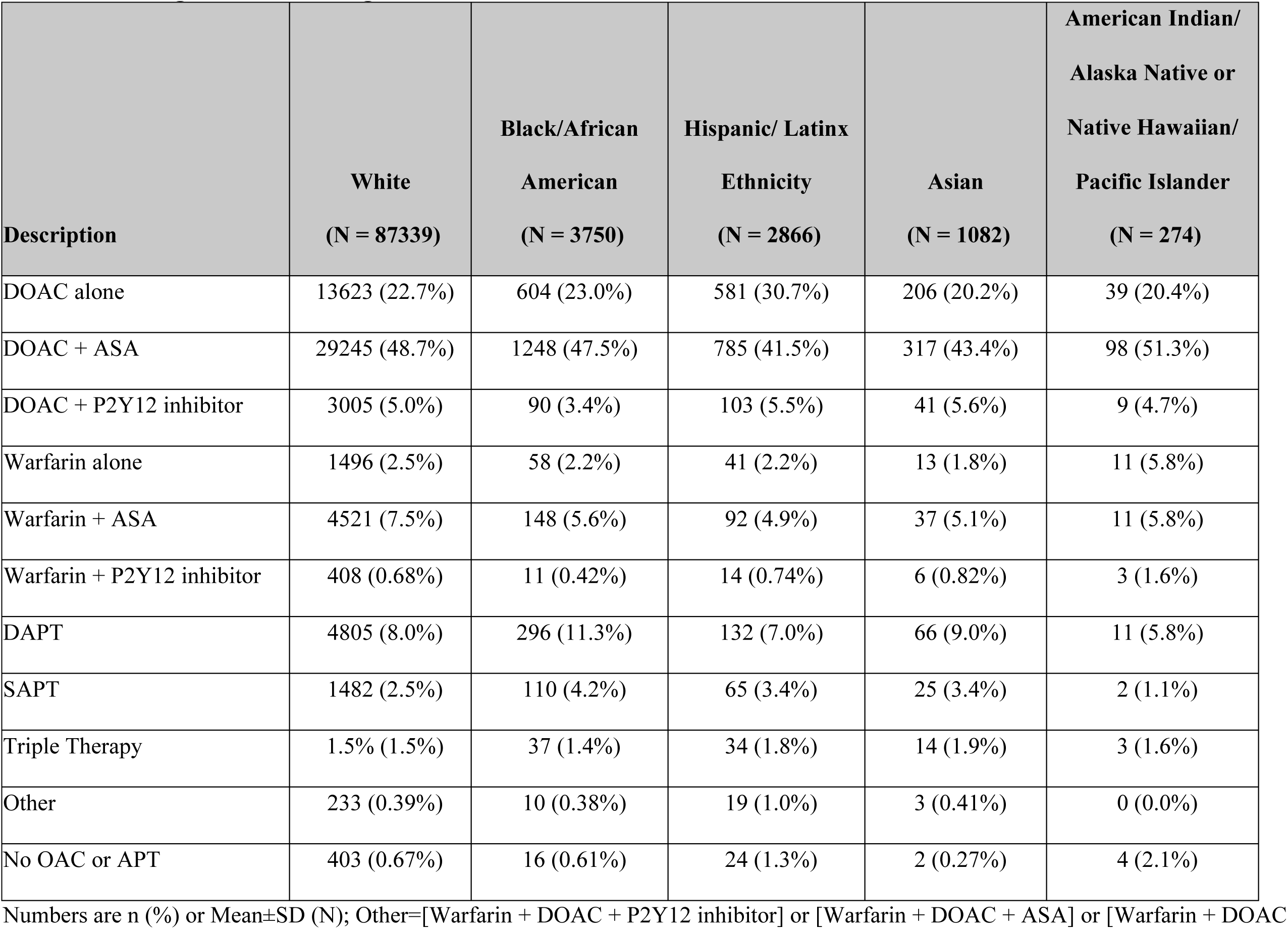
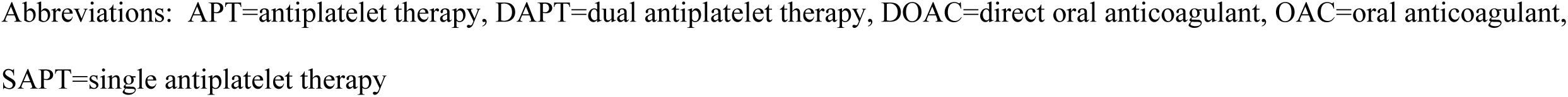
Discharge Medication Regimens.

| **Description** | **White**<br>**(N = 87339)** | **Black/African**<br>**American**<br>**(N = 3750)** | **Hispanic/ Latinx**<br>**Ethnicity**<br>**(N = 2866)** | **Asian**<br>**(N = 1082)** | **American Indian/**<br>**Alaska Native or**<br>**Native Hawaiian/**<br>**Pacific Islander**<br>**(N = 274)** |
| --- | --- | --- | --- | --- | --- |
| DOAC alone | 13623 (22.7%) | 604 (23.0%) | 581 (30.7%) | 206 (20.2%) | 39 (20.4%) |
| DOAC + ASA | 29245 (48.7%) | 1248 (47.5%) | 785 (41.5%) | 317 (43.4%) | 98 (51.3%) |
| DOAC + P2Y12 inhibitor | 3005 (5.0%) | 90 (3.4%) | 103 (5.5%) | 41 (5.6%) | 9 (4.7%) |
| Warfarin alone | 1496 (2.5%) | 58 (2.2%) | 41 (2.2%) | 13 (1.8%) | 11 (5.8%) |
| Warfarin + ASA | 4521 (7.5%) | 148 (5.6%) | 92 (4.9%) | 37 (5.1%) | 11 (5.8%) |
| Warfarin + P2Y12 inhibitor | 408 (0.68%) | 11 (0.42%) | 14 (0.74%) | 6 (0.82%) | 3 (1.6%) |
| DAPT | 4805 (8.0%) | 296 (11.3%) | 132 (7.0%) | 66 (9.0%) | 11 (5.8%) |
| SAPT | 1482 (2.5%) | 110 (4.2%) | 65 (3.4%) | 25 (3.4%) | 2 (1.1%) |
| Triple Therapy | 1.5% (1.5%) | 37 (1.4%) | 34 (1.8%) | 14 (1.9%) | 3 (1.6%) |
| Other | 233 (0.39%) | 10 (0.38%) | 19 (1.0%) | 3 (0.41%) | 0 (0.0%) |
| No OAC or APT | 403 (0.67%) | 16 (0.61%) | 24 (1.3%) | 2 (0.27%) | 4 (2.1%) |

Numbers are n (%) or Mean±SD (N); Other=[Warfarin + DOAC + P2Y12 inhibitor] or [Warfarin + DOAC + ASA] or [Warfarin + DOAC
Abbreviations: APT=antiplatelet therapy, DAPT=dual antiplatelet therapy, DOAC=direct oral anticoagulant, OAC=oral anticoagulant, SAPT=single antiplatelet therapy

### Outcomes at 45 days

Outcomes at 45 days, unadjusted for differences in baseline characteristics, are shown in **Figure 1** and **Supplemental Table 4**. Death occurred in 0.81% of patients overall at 45 days and was not statistically different across racial and ethnic groups ranging from 0.41% in Native American/Pacific Islanders to 1.10% in Black patients. Stroke occurred in 0.29% of patients overall and was highest in Asian patients (0.80%) at 45 days. The rate of major bleeding in all patients combined was 3.11% at 45 days. There were significant statistical differences overall, the highest rates were found in Black patients (4.92%) and Native American/Pacific Islanders (4.89%). Peridevice leak at 45 days was similar across groups.

**Figure 1.**
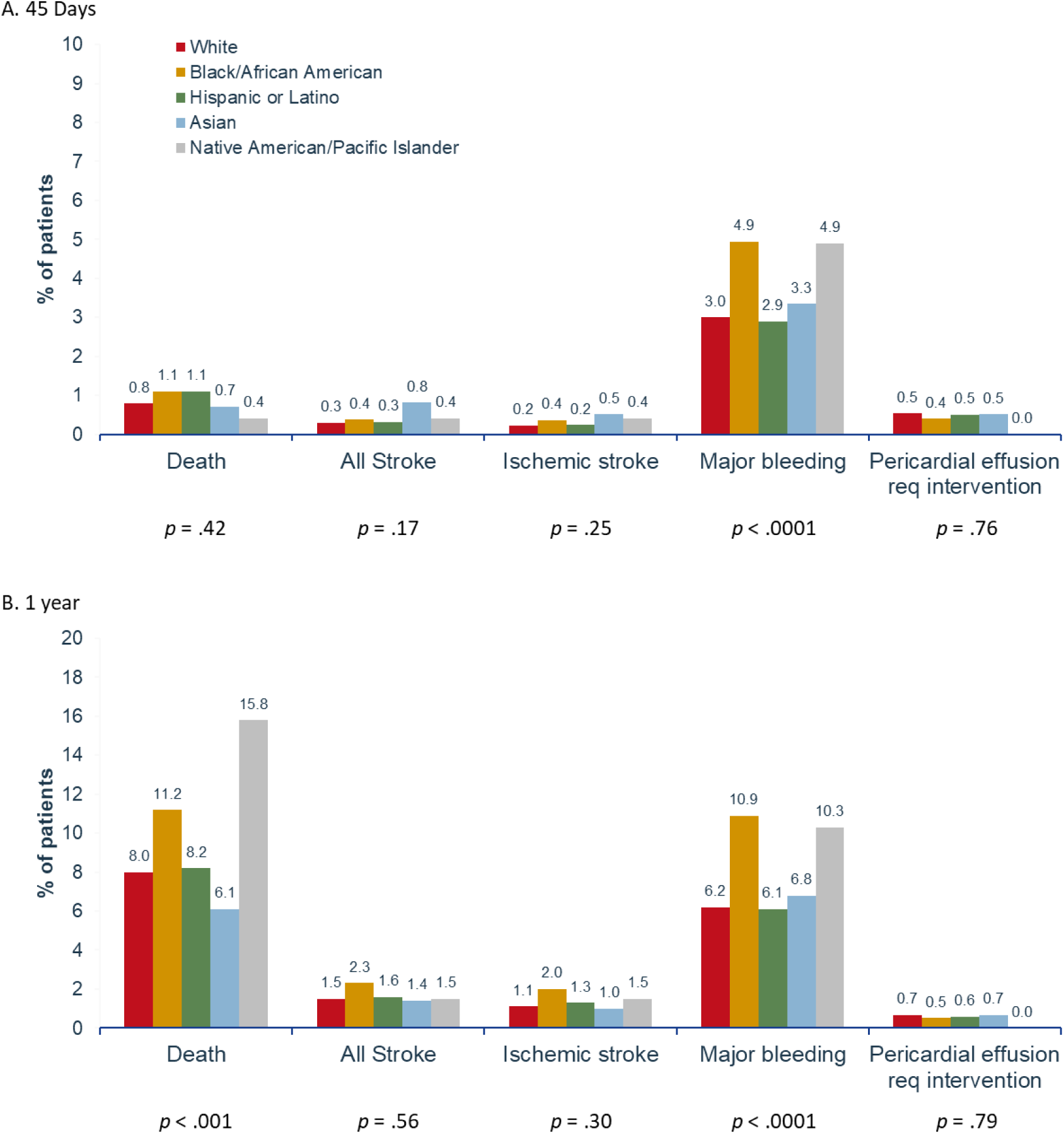
Outcomes at 45 day and 1 year by race and ethnicity. Time-to-event outcomes at (A) 45 days and (B) 1 year by race and ethnicity in patients implanted with WATCHMAN FLX.

Cox proportional hazard regression analysis to adjust for differences in baseline characteristics, was performed in the three largest racial and ethnic groups using White patients as the reference group (**Table 4**, **Figure 2**). After adjustment, the risk of bleeding was significantly higher at 45 days in Black compared to White patients (HR 1.30, 95% confidence interval 1.10, 1.54). Death and stroke were similar between Black and White patients after adjustment. No differences in death, stroke or bleeding were found in White vs Hispanic patients at 45 days after adjustment.

**Figure 2.**
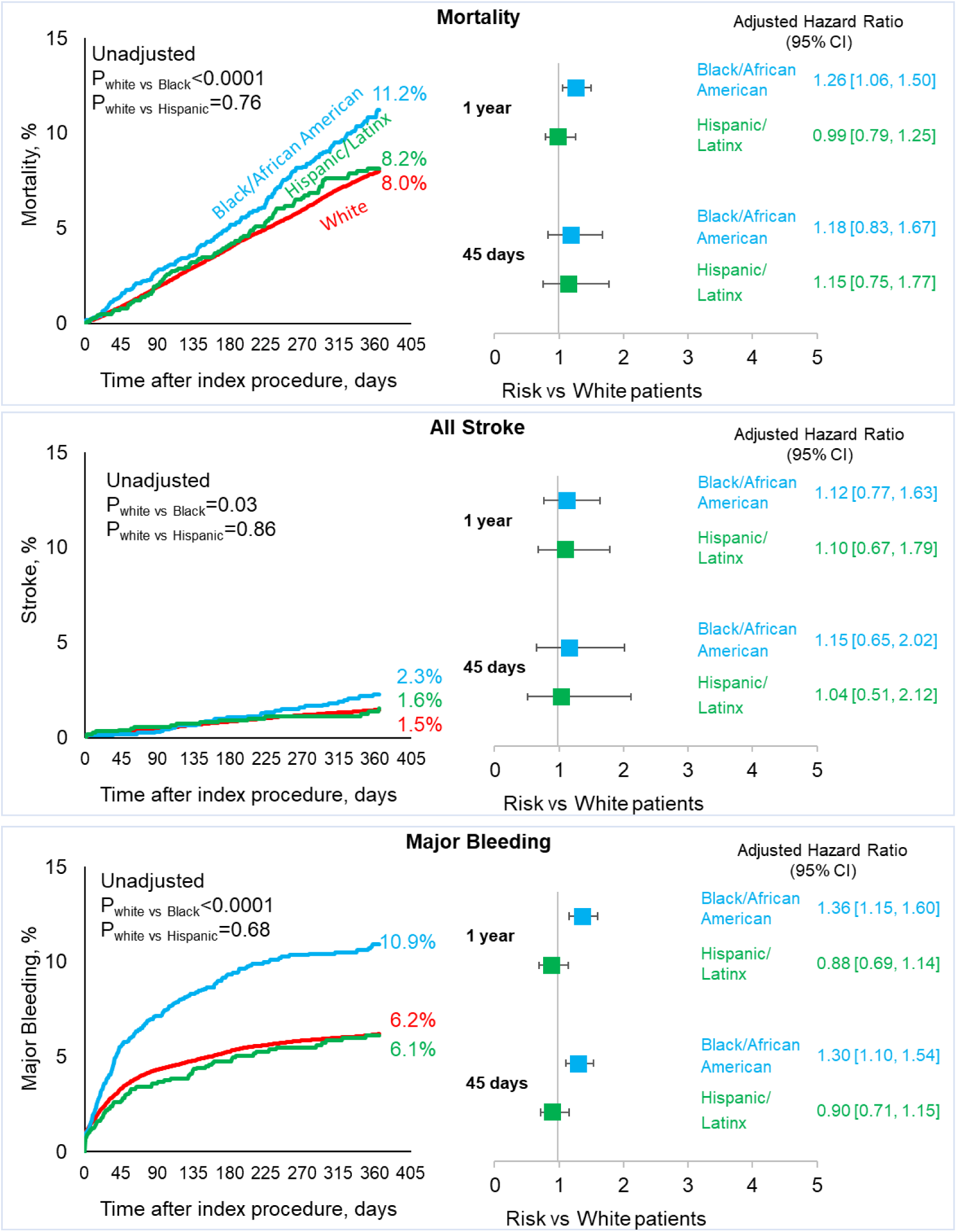
Unadjusted and Adjusted Outcomes of Adverse Events for White, Black/African American, and Hispanic/Latinx Patients Receiving WATCHMAN FLX. P-value from the log-rank test. Hazard ratio (HR) [95% confidence interval]; reference White patients; *Major bleeding defined as bleeding requiring hospitalization, and/or causing a decrease in hemoglobin level > 2g/dL, and/or requiring blood transfusion that was not hemorrhagic stroke. Multivariable models adjusted for age, gender, CHA_2_DS_2_-VASc and HAS-BLED score components, type of atrial fibrillation, diabetes, fall risk, history of bleeding, chronic lung disease, sleep apnea, cardiomyopathy, coronary artery disease, prior ablation, left ventricular ejection fraction and post implant device margin residual leak.

**Table 4.** Outcomes in the adjusted cohorts through 45 day and 1 year follow-up visits.

|  | Black/African American |  | Hispanic/Latinx Ethnicity |  |
| --- | --- | --- | --- | --- |
|  | Univariate<br>HR [95% CI] | Multivariate<br>HR [95% CI] | Univariate<br>HR [95% CI] | Multivariate<br>HR [95% CI] |
| **45 days** |  |  |  |  |
| All Death | 1.39 [1.00, 1.94] | 1.18 [0.83, 1.67] | 1.11 [0.72, 1.70] | 1.15 [0.75, 1.77] |
| All Stroke | 1.30 [0.74, 2.28] | 1.15 [0.65, 2.02] | 1.10 [0.54, 2.23] | 1.04 [0.51, 2.12] |
| Ischemic Stroke | 1.69 [0.96, 2.98] | 1.50 [0.85, 2.64] | 1.07 [0.48, 2.43] | 1.06 [0.47, 2.40] |
| Major Bleeding* | 1.64 [1.39, 1.92] | 1.30 [1.10, 1.54] | 0.99 [0.79, 1.25] | 0.90 [0.71, 1.15] |
| **1 year** |  |  |  |  |
| All Death | 1.37 [1.17, 1.61] | 1.26 [1.06, 1.50] | 1.04 [0.83, 1.29] | 0.99 [0.79, 1.25] |
| All Stroke | 1.38 [0.97, 1.96] | 1.12 [0.77, 1.63] | 1.06 [0.65, 1.73] | 1.09 [0.67, 1.79] |
| Ischemic Stroke | 1.57 [1.08, 2.28] | 1.24 [0.83, 1.85] | 1.12 [0.66, 1.92] | 1.13 [0.66, 1.94] |
| Major Bleeding* | 1.75 [1.50, 2.04] | 1.36 [1.15, 1.60] | 0.99 [0.78, 1.25] | 0.89 [0.69, 1.14] |

### Outcomes at 1 year

Outcomes at 1 year are show in **Figure 1** and **2**, **Supplemental Table 6**. At 1 year, the rate of unadjusted mortality was 8.15% and was significantly different across the racial and ethnic groups. Native American/Pacific Islanders (15.79%) and Black (11.24%) had the highest rates of death at 1 year. Major bleeding in the overall patient cohort was 6.37% at 1 year and varied significantly among racial and ethnic groups. Major bleeding was highest in Native American/Pacific Islanders (10.32%) and Black (10.91%) patients compared to the other groups (**Figure 1** and **2**, **Supplemental Table 6**). Stroke overall at 1 year was 1.48% and, though not statistically significantly different, ranged from 1.37% in Asian patients to 2.28% in Black patients.

After adjustment, the risk of death and bleeding was significantly higher in Black compared to White patients at 1 year (death: 1.26 [1.06, 1.50]; bleeding: HR 1.36 [1.15, 1.60]). Hispanic and White patients had similar risk of death and bleeding at 1 year. The risk of stroke was similar between groups in each comparison (White vs Black or White vs Hispanic) after adjustment at 1 year.

## Discussion

This study is the largest analysis of racial and ethnic differences in utilization and outcomes of LAAO procedures in the US to date including almost 100,000 patients. We demonstrated the following key findings: 1) significant racial and ethnic disparities exist, less than 4% Black and less than 3% and Hispanic patients comprise patients receiving LAAO ; 2) Black patients undergoing LAAO are significantly younger but with more comorbidities than White patients undergoing the procedure; and 3) the risk of death at 1 year and major bleeding at 45 days and 1 year was higher for Black patients compared to White patients.

### Disparities in Access to LAAO

The racial and ethnic representation among WATCHMAN FLX recipients in this study mirrors the low enrollment of racial and ethnic minority patients in clinical trials involving these devices. The PROTECT-AF and PREVAIL trials that that led to US FDA approval for the WATCHMAN device had very few minority patients with 1.3% and 2.2% Black enrollment, respectively, as compared to >90% White enrollment in both trials.^15,16^ Similar disparities in real world LAAO utilization have been previously described in other observational studies.^10,17–19^ An important consideration that affects access of racial and ethnic minority patients to LAAO procedures is the location of hospitals/centers that perform the procedures. An analysis of Medicare fee-for-service claims (2016 to 2019) by Reddy and colleagues revealed that between 2016 and 2019, 97.4% of new LAAO programs opened in the US were in metropolitan areas.^7^ The authors found that LAAO centers, as compared to non-LAAO centers, provided care to patients with higher median household incomes. After adjustment for socioeconomic markers, age, and clinical comorbidities, LAAO procedures were less likely to be performed in zip codes with higher proportions of Black and Hispanic patients.^7^ Another important consideration that affects the demographic composition of those who undergo LAAO procedures is that White patients have a higher risk of AF than other racial and ethnic groups.^4^ However, this only explains some of the underrepresentation of racial and ethnic minority groups among LAAO patients. In one analysis using the National Inpatient Sample (NIS, a 20% random, stratified sample of hospital discharges in the US), there were racial and ethnic differences in adult admissions related to a diagnosis of AF where more than 80% were White, 9.3% were Black, 5.7% were Hispanic, 2% were Asian American and Pacific Islander, and 2.4% were of other races and ethnicities.^18^ However, the proportions of racial and ethnic minority patients undergoing LAAO procedures was still lower than their proportions among AF admissions. Examining the racial and ethnic make-up of the patients who underwent LAAO in this study, Black patients made up only 4.2% and Hispanic patients 5.0% of LAAO patients.^18^ Similar underrepresentation was seen in other NIS analyses.^10,17,19^

### Disparities in LAAO Outcomes

Prior analyses have focused on short-term outcomes and found differences in major adverse events between White patients and racial and ethnic minority patients regarding intra-procedural or immediate postprocedural complications even after adjusting for differences in baseline characteristics.^10,17–19^ SURPASS patients were followed up to a year and the rates of major adverse events at 45 days and 1 year were similar in White and Hispanic patients. However, the risk of death at 1 year and major bleeding at 45 days and 1 year was higher for Black patients compared to White patients even after adjustment for differences in baseline comorbidities.

The differences in outcomes in this study may be related to differences in medical comorbidities. Black and Hispanic patients had higher CHA_2_DS_2_-VASc and HAS-BLED scores. Additionally, Black patients had the highest incidence of prior bleeding and more than twice the incidence of abnormal renal function compared to White patients. This increase in bleeding in Black patients may be driven by worse renal function in this patient population. These data echo other studies focusing on the impact of race and ethnicity after LAAO. Compared with White patients, more Black patients, Hispanic patients, and patients of Asian, Asian American, or Pacific Islander ancestry had diabetes mellitus, chronic kidney disease, previous stroke, and deficiency anemias.^18^

Black patients were more commonly discharged on anticoagulation plus aspirin or DAPT instead of anticoagulation alone, which has been previously been reported to be associated with higher rates of bleeding after LAAO.^20^ The increased use of antiplatelet agents may be indicated in this population with a higher burden of cardiovascular and renal comorbidities but could impact the difference in outcomes that were found between White and Black patients. Residual confounding due to unreported patient characteristics, disparities in access to LAAO, and post-procedural care may also account for some of the differences in bleeding and mortality observed.

### Future Directions

Disparities leading to inadequate access to structural heart interventions remains a challenge in other areas as well, as similar single digit representation of African American and Hispanic patients were found in the TAVR population.^5,9,21^ An analysis of the Transcatheter Valve Therapy Registry of patients undergoing TAVR in the US revealed only 3.8% of patients were African American compared to 91.3% White patients.^9^ Similar findings have been found for other cardiovascular procedures including mitral valve replacement, and implantation of implantable cardioverter-defibrillators.^6,8,9^ There is a pressing need to increase enrollment of minority patients into clinical trials either by way of mandated enrollment targets, broadening the clinical trial sites to include hospitals serving predominantly racial and ethnic minoritized patients and including investigators who also serve this patient population in clinical trials.

Strategies to improve access should also include efforts to train providers and help develop infrastructure at centers that provide care to mainly minority patients. Interestingly, hospitals in urban centers with a large minority population still have low treatment rates of minority patients^7^, suggesting that low enrollment rates are not just a location issue. Efforts must be made to address barriers to care even in hospitals serving predominantly minority communities. Even if trial enrollment of minority populations improves, the absolute numbers of minority patients may remain relatively modest, particularly for device trials which tend to be smaller, precluding detailed assessments of differences in outcomes. Real-world registry data serves as a complement to smaller trials for the study of these important patient subgroups.

### Limitations

As with all observational registries there are limitations to this analysis including unmeasured or residual confounders. Both race and ethnicity were defined by the subject but are social constructs that may encompass a variety of genetic and cultural backgrounds. Cancelled procedures are not included in the data received from the NCDR LAAO registry and hospital location or region are not known. Dialysis status is unknown and adjusting for eGFR stage may not capture different degrees of abnormal renal function. There is no control group, so it is difficult to determine if disparities are inherent to differences in efficacy versus differences in baseline risk. Only patients treated with WATCHMAN FLX are included and patients are being followed for more than 2 years but this analysis is limited to 1 year follow-up. The reporting of events relies on site-reported data although algorithmic adjudication of adverse events is employed by the NCDR LAAO registry.

## Conclusions

This study is the largest analysis of racial and ethnic disparities in LAAO procedures in the US to date and it revealed that minorities remain significantly under-represented despite increasing procedural volumes nationally. Black patients were younger but had higher baseline comorbidities and experienced higher rates of bleeding at 45 days and 1 year and higher 1-year mortality

## Data Availability

All data is available

## Acknowledgements

Funding/Support

This analysis was supported by the Boston Scientific Corporation. The authors thank and Kristine Roy, PhD for writing/editing assistance, and Yanrong Zhu, PhD for assistance with statistical analysis (paid employees of Boston Scientific Corporation).

## Abbreviations list

ACC: American College of Cardiology
LAAO: left atrial appendage occlusion
NCDR: National Cardiovascular Data Registry
AF: atrial fibrillation
OAC: oral anticoagulation
DAPT: dual antiplatelet therapy
DOAC: direct oral anticoagulant

